# Genomic diversity of SARS-CoV-2 in Pakistan during fourth wave of pandemic

**DOI:** 10.1101/2021.09.30.21264343

**Authors:** Massab Umair, Aamer Ikram, Zaira Rehman, Adnan Haider, Nazish Badar, Muhammad Ammar, Abdul Ahad, Rana Suleman, Muhammad Salman

**Author notes:** **Corresponding author:** Massab Umair, Department of Virology, National Institute of Health, 45500, Park Road, Chak Shahzad, Islamabad, Pakistan.

## Abstract

The emergence of different variants of concern of SARS-CoV-2 has resulted in upsurges of COVID positive cases around the globe. Pakistan is also experiencing fourth wave of COVID-19 with increasing number of positive cases. In order to understand the genomic diversity of circulating SARS-CoV-2 strains during fourth wave of pandemic in Pakistan, the current study was designed. The samples from 89 COVID-19 positive patients were subjected to whole genome sequencing using GeneStudio S5. The results showed that 99% (n=88) of isolates belonged to delta variant and only one isolate belonged to alpha variant. Among delta variant cases 26.1% (n=23) isolates were showing B.1.617.2 while 74% of isolates showing AY.4 lineage. Islamabad was found to be the most affected city with 54% (n=48) of cases, followed by Karachi (28%, n=25), and Rawalpindi (10%, n=9). AY.4 has slight difference in mutation profile compared to B.1.617.2. E156del, G142D and V26I mutations in spike and T181I in NSP6 were present in B.1.617.2 but not in AY.4. Interestingly, A446V mutation in NSP4 has been only observed in AY.4. The current study highlights the circulation of primarily delta variant (B.1.617.2 and AY.4) during fourth wave of pandemic in Pakistan.

## Introduction

Severe Acute Respiratory Syndrome Coronavirus 2 (SARS-CoV-2) belongs to family coronaviridae that cause moderate to severe respiratory illness. As of September 27, 2021, 232,612,493 people have been infected worldwide with 4,726,175 fatalities [1]. Since the commencement of the COVID-19 pandemic, different mutations have emerged that ultimately lead to various lineages of SARS-CoV-2 [2]. These lineages were then classified as Variants of Concern (VOCs) and Variants of Interest (VOIs). There are currently four VOCs (Alpha, Beta, Gamma, and Delta), two VOIs (Lambda and Mu) and four variants are under investigation (Eta, Theta, Iota, and Kappa) [3]. Among the VOCs, delta was found to be the highly transmissible variant after alpha and first identified from India in September 2020. It is currently the second most prevalent VOC, which is now spread in more than 100 countries[4]. In Pakistan the first case of delta variant was detected on May 16, 2021. It has been reported that delta variant is 60% more transmissible than alpha. It also has the potential for reduction in neutralization by some EUA monoclonal antibody treatments and post-vaccination sera [5]. The Pfizer-BioNTech vaccine appeared to give 79% protection against delta variant infection after 14 days of getting a second dose, compared to 92% protection against the alpha variant [6].

In Pakistan, 1,250,425 individuals have been infected with 27,597 deaths as of September 29, 2021 [1]. Pakistan has experienced three waves of COVID-19 pandemic and currently is under the fourth wave. The first wave was from May-July 2020, the second was from October, 2020-January, 2021, the third wave began in March, 2021 and ended in May, 2021 while the fourth wave begins in July 2021 [7]. The average positivity rate in the start of fourth wave was 25% and now drops to 5% (as of September, 29, 2021). Despite such a high positivity rate during the fourth wave, the data regarding the circulating strains of SARS-CoV-2 is not available. Hence, in order to explore the genomic diversity of SARS-CoV-2 during fourth wave of pandemic in Pakistan whole genome sequencing of SARS-CoV-2 positive samples were performed.

## Materials and Methods

### Sample Collection

The oropharyngeal swab specimens of subjects who tested positive during clinical RT-PCR testing were retrospectively collected (n=89) and used in the present analysis. The study was approved by ethical review committee of National Institute of Health, Islamabad.

### RNA Extraction and real-time PCR

Total RNA was extracted from the specimens using KingFisher™ Flex Purification System (ThermoFisher Scientific, US). Clinical RT-PCR testing was performed using TaqPath™ COVID-19 CE-IVD RT-PCR kit (ThermoFisher Scientific, Waltham, US) for SARS-CoV-2 at National Institute of Health, Islamabad, Pakistan. The COVID-19 positive specimens with low cycle threshold (Ct) values (≤ 27) followed by selection based on their geographical location, i.e., representing the entire country were selected for whole genome sequencing.

### cDNA synthesis, and Next Generation Sequencing

The cDNA synthesis was performed using SuperScript™ IV VILO™ Master Mix (Invitrogen, USA). The library was prepared using the Ion Ampliseq™ Library Kit Plus (Thermofisher Scientific, US) and Ion AmpliSeq™ SARS-CoV-2 research assay panel (Thermofisher Scientific, US) according to the manufacturer’s instructions. The prepared library underwent template preparation and enrichment with the Ion Chef system. The enriched templates were then loaded onto an Ion 540 chip for semiconductor sequencing on the Ion GeneStudio S5 machine using the Ion S5 sequencing reagents (Thermofisher Scientific, US) as per manufacturer’s protocols.

### Data Analysis

The Fastq files were exported using File Exporter plugin from Torrent Suite 5.14.0 and then processed for quality assessment using the FastQC tool (v0.11.9) [8]. Trimmomatic (v0.39) [9] was employed to eliminate artifacts and technical biases by removing adapter sequences and low-quality base calls (< 20). The filtered reads were aligned using the Burrows-Wheeler Aligner’s (BWA, v0.7.17) [10] and available reference genome (Wuhan-Hu-1, Accession number: MN908947.3). According to Centers for Disease Control and Prevention (CDC, USA) guidelines, variants were identified and consensus sequences for all genomes were generated [11]. Pangolin v3.1.11 and pangoLEARN model dated 09-08-2021 were used to classify the assembled genomes into PANGO lineages [12].

## Results

During July 19, 2021 to August 24, 2021 a total of 89 whole genomes of SARS-CoV-2 were sequenced at National Institute of Health, Islamabad. The age range of patients were 3-90 years with median age of 32 years. The male to female ratio was 53:36. Out of the 89 isolates, 88 were delta variant and one was alpha variant. Among the delta variant cases, 23 isolates belonged to B.1.617.2 and interestingly, 65 isolates belonged to AY.4 (sub-lineage of B.1.617.2) and one isolate belonged to B.1.1.7 (alpha). According to geographical distribution, 54% (n=48) of cases were reported from Islamabad, followed by Karachi (28%, n=25), and Rawalpindi (10%, n=9). Among the AY.4 cases (n=65), 52% (n=34) of isolates were from Islamabad, 24.6% (n=16) from Karachi and 13.8% (n=9) from Rawalpindi. One case of AY.4 were reported each from Peshawar, Sargodha, Azad Kashmir and Panjgur (Figure 1).

**Figure 1:**
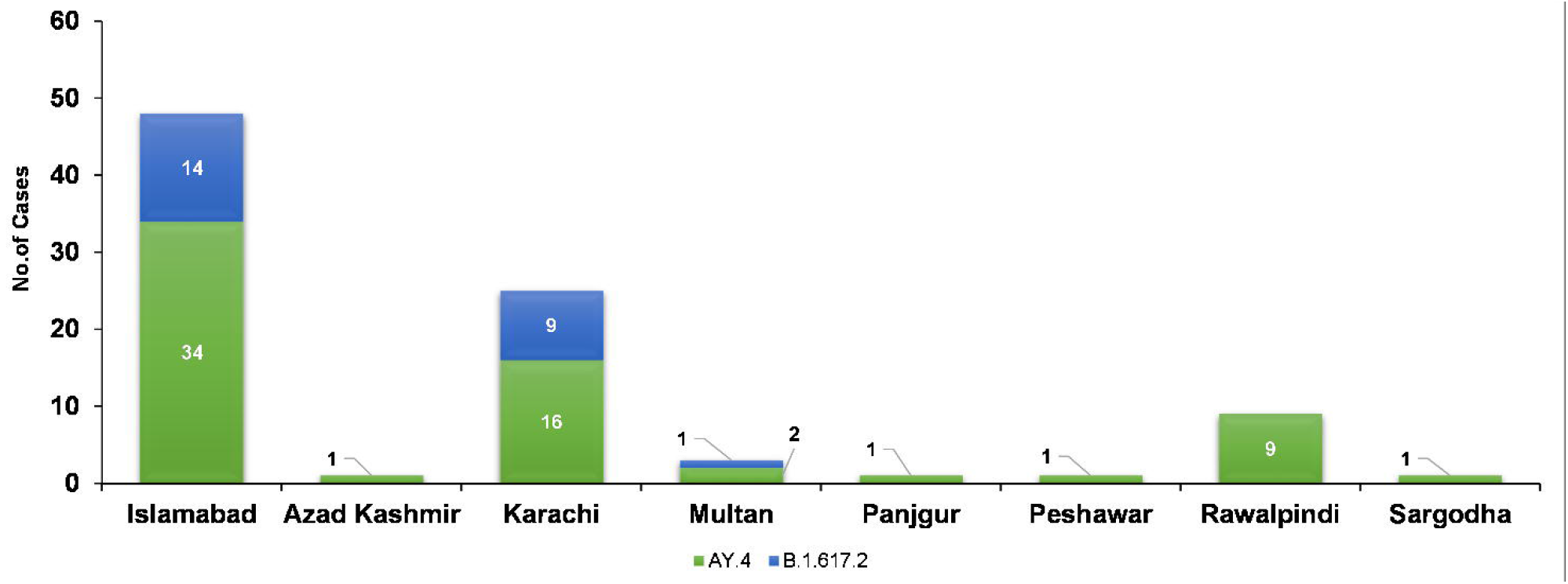
Geographical distribution of delta variant cases (B.1.617.2, and AY.4).

We have found a total of 428 nucleotide changes in 89 studied genomes (Figure 2). Out of these 428, 253 have been the non-synonymous single nucleotide polymorphisms (SNPs) and 168 have been synonymous SNPs. In addition these sequences harbors 04 deletions and 3 stop codons. Among the non-synonymous SNPs, there have been 42 SNPs with >2% frequency (Figure 3).

**Figure 2:**
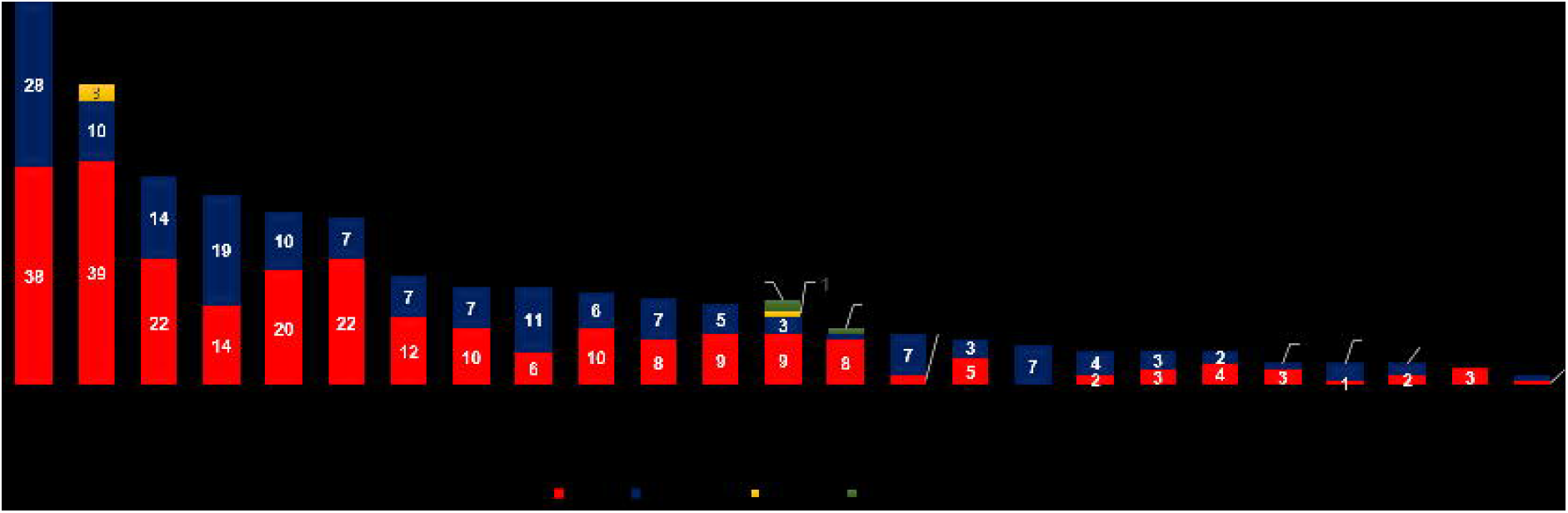
Total number of mutation identified in 89 isolates.

**Figure 3:**
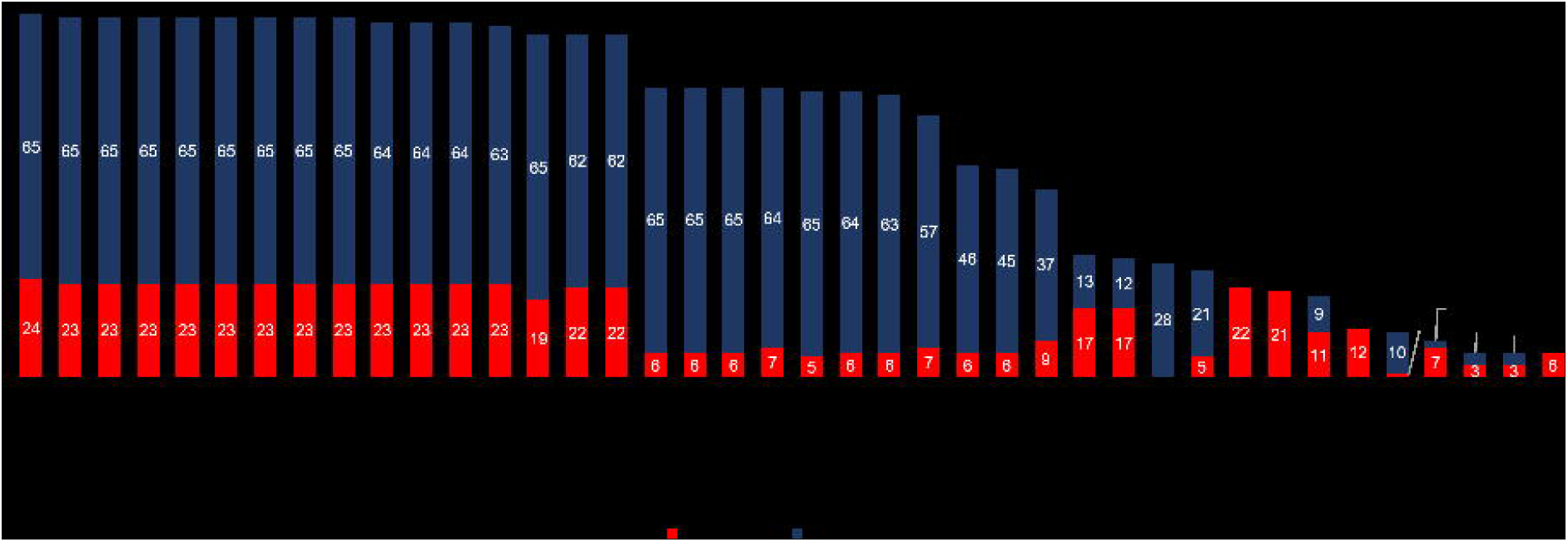
Number of single-nucleotide mutation (occurred in more than 2% of the samples) present in different genomic segments of SARS-CoV-2 genome

Observably, the ORF1ab has been the most mutated protein with highest number of mutations in NSP3 and NSP4 while NSP1-2, NSP5, NSP7-11 and NSP-15 have been relatively conserved. While comparing the mutation profile of B.1.617.2 and AY.4, we have found similar characteristics of spike mutations (L452R, P681R, T19R, T478K, and D950N) in both the lineages In ORF7a, a stop codon has been observed at position 94 as Q94* (EPI_ISL_3827532; AY.4). However, in AY.4 lineage some novel mutations have been identified as A446V in NSP4 while in case of S protein the E156del, G142D, and V26I have been present in B.1.617.2 and not in AY.4. Another mutation (T81I) in NSP6 have been found in B.1.617.2 and not in AY.4 (Table 1). In addition, we have also identified some unique mutations in the studied sequences. In one of the isolate an unusual stop codon have been observed in ORF8 at position 68 (K68*) which belonged to alpha variant. In the AY.4 lineage some unique mutations have been identified in spike glycoprotein as A845S (GISAID ID: EPI_ISL_3462513), G446V (GISAID ID: EPI_ISL_3462511), N532S (GISAID ID: EPI_ISL_3553517; AY.4) and V483A/F (AY.4). In case of alpha variant only one unique spike mutation is observed as K529N (GISAID ID: EPI_ISL_3553513).

**Table 1:**
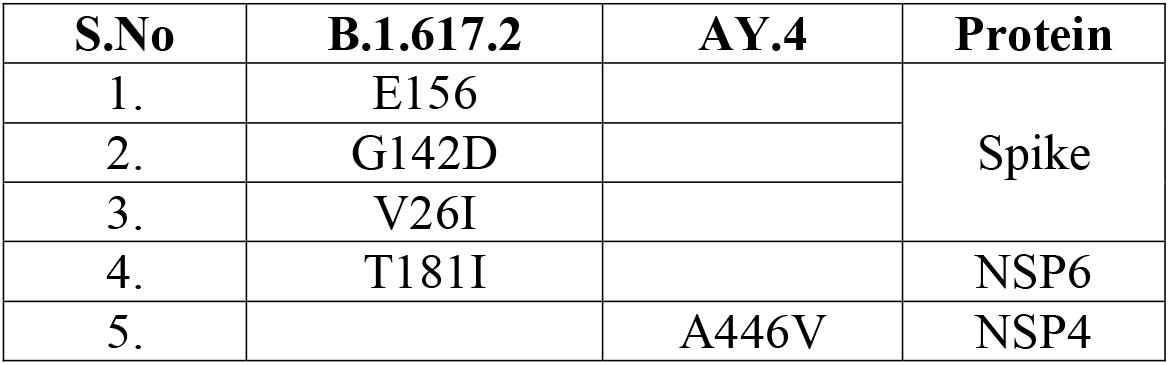
Comparison of B1.617.2 and AY.4 in terms of amino acid residues.

## Discussion

Delta variant of SARS-CoV-2 is now becoming the most dominant variant globally with spread into more than 100 countries from UK, USA, Asia, and Africa. It is one the most transmissible variant among other variants of SARS-CoV-2. Since the detection of first case of delta variant from Pakistan the number of positive cases start increasing and Pakistan experience fourth wave starting from mid of July, 2021. In-order to understand the circulating strain during the fourth wave current study was designed. We have identified an upsurge of AY.4 cases in Pakistani population. This data is also supported by the global distribution of delta variant and its lineages. According to sequencing data reported globally, AY.4 is one the most prevalent lineage after B.1.617.2 [13]. Globally, the upsurge of AY.4 was observed since June, 2021 and it is now dominating in the world. The overall prevalence of delta variant around the globe is 38% with B1.617.2 having prevalence of 12% and AY.4 is 17% prevalent (as of September 28, 2021). The data reported in the current study is in accordance with the worldwide stats where 73% of isolates belonged to AY.4. Islamabad was found to be having the highest number of AY.4 cases (52% of all the AY.4 cases) followed by Karachi and Rawalpindi. The highest positivity rate in Islamabad (10%), Karachi (23%) and Rawalpindi (18%) in the last week of July, 2021 was attributed to the circulation of delta variant in population.

At the sequence level there is a slight difference in the mutation profile of B.1.617.2 and AY.4. The AY.4 isolates does not have V26I, G142D, and E156del mutations of spike protein that are found in B.1.617.2. In the non-structural protein 6 (NSP6) T181I was not observed in any of the AY.4 isolates. Interestingly, one mutation identified in the NSP4 region as A446V and is not found in any of the B.1.617.2 isolates.

This study highlights the circulations of delta variant (B.1.617.2 and AY.4) during the fourth wave of pandemic in Pakistan. The highest number of cases during fourth wave also supported by the high transmissibility ratio of delta variant. But the effective vaccination campaign resulted in less severity during fourth wave as observed in India. As of September 28, 2021, 36.7% of the population received either first or second dose of vaccine. Despite the high transmissibility rate vaccination lowers the disease severity. It has been documented that in countries where the vaccination rate is <15% like Thailand, South Africa, and Indonesia, delta has ripped through cities and threatens to overwhelm healthcare infrastructure. The sudden surge of delta variant cases in India was also due to slow vaccination rate. A study by Public Health England (PHE) states that although the delta variant makes the vaccine less effective but still prevent hospitalization in more than 90% of cases [14]. The same observed in Pakistan, where we have increased number of cases but have less deaths and hospitalization with case fatality rate of 2%.

The dynamics of 4^th^ wave of COVID-19 pandemic in Pakistan has shown interesting epidemic modifying pattern which holds importance in national decision making for handling the pandemic. The decrease in hospitalization even with a highly transmissible variant of concern AY.4 suggest that vaccination coverage might help in suppressing future waves of the COVID-19 in Pakistani population. Furthermore, the disease also highlights the continued effectiveness of genomic surveillance of SARS CoV2 variants.

## Data Availability

All the sequences generated in the current study are submitted to the GISAID that are available at https://www.gisaid.org/login/ under the accession numbers: EPI_ISL_3462498- EPI_ISL_3462527, EPI_ISL_3553499-EPI_ISL_3553528, EPI_ISL_3827529-EPI_ISL_3827557.

## Conflict of Interest

All the authors declared no conflict of interest.

## Data Availability

All the sequences generated in the current study are submitted to the GISAID that are available at “https://www.gisaid.org/login/” under the accession numbers: EPI_ISL_3462498-EPI_ISL_3462527, EPI_ISL_3553499-EPI_ISL_3553528, EPI_ISL_3827529-EPI_ISL_3827557.

## Notes

### Competing Interest Statement

The authors have declared no competing interest.

### Funding Statement

No funding received for the study.

### Author Declarations

The study was approved by ethical review committee of National Institute of Health.

## REFERENCES

1. “COVID Live Update: 233,105,342 Cases and 4,769,835 Deaths from the Coronavirus - Worldometer.” [Online]. Available: https://www.worldometers.info/coronavirus/. [Accessed: 28-Sep-2021].

2. E. Cella et al., “SARS-CoV-2 Lineages and Sub-Lineages Circulating Worldwide: A Dynamic Overview,” Chemotherapy, vol. 66, no. 1–2, p. 1, Jun. 2021.

3. Julia L. Mullen et al., “outbreak.info.” [Online]. Available: https://outbreak.info/citation. [Accessed: 28-Sep-2021].

4. A. Vaughan, “Delta to dominate world,” New Sci., vol. 250, no. 3341, p. 9, Jul. 2021.

5. CenterDC, “SARS-CoV-2 Variant Classifications and Definitions,” Cdc, 2021. [Online]. Available: https://www.cdc.gov/coronavirus/2019-ncov/variants/variant-info.html.

6. Linda Geddes, “Five things we know about the Delta variant (and two things we don’t) | Gavi, the Vaccine Alliance,” 2021. [Online]. Available: https://www.gavi.org/vaccineswork/five-things-we-know-about-delta-coronavirus-variant-and-two-things-we-still-need. [Accessed: 28-Sep-2021].

7. “COVID-19 Health Advisory Platform by Ministry of National Health Services Regulations and Coordination.” [Online]. Available: https://covid.gov.pk/. [Accessed: 28-Sep-2021].

8. Andrews, S., FastQC: a quality control tool for high throughput sequence data. 2010, Babraham Bioinformatics, Babraham Institute, Cambridge, United Kingdom.

9. Bolger, A.M., M. Lohse, and B. Usadel, Trimmomatic: a flexible trimmer for Illumina sequence data. Bioinformatics, 2014. 30(15): p. 2114–2120.

10. Li, H. and R. Durbin, Fast and accurate long-read alignment with Burrows–Wheeler transform. Bioinformatics, 2010. 26(5): p. 589–595.

11. Paden, C.R., et al., Rapid, sensitive, full-genome sequencing of severe acute respiratory syndrome coronavirus 2, https://www.nc.cdc.gov/eid/article/26/10/20-1800_article. Emerging infectious diseases, 2020. 26(10): p. 2401.

12. O’Toole, A., et al., Pangolin: lineage assignment in an emerging pandemic as an epidemiological tool. 2020.

13. https://outbreak.info/situation-reports/delta?loc=IND&loc=GBR&loc=USA&selected

14. https://qz.com/india/2036052/delta-variant-is-causing-a-surge-in-covid-cases-globally/

